# Genomic surveillance of SARS-CoV-2 reveals emergence of Omicron BA.2 in Islamabad, Pakistan

**DOI:** 10.1101/2022.02.23.22271372

**Authors:** Massab Umair, Aamer Ikram, Zaira Rehman, Syed Adnan Haider, Muhammad Ammar, Nazish Badar, Qasim Ali, Muhammad Suleman Rana, Muhammad Salman

## Abstract

The Omicron variant of SARS-CoV-2 has rapidly replace previous variants of SARS CoV2 around the globe and is now a major variant of concern. The genomic surveillance of Omicron variant also reveals spread of its subvariant BA.2 which has differing transmissibility in comparison to its other subvariant BA.1. BA.1 and BA.2 harbors different mutational profile. One of the important change in both the subvariants is the presence of 69-70 deletion in BA.1 and absence of this deletion in BA.2. This deletion can be used as tool for the detection of omicron sub variants using real time PCR. In the current study we have used the TaqPath COVID-19 PCR kit for the detection of 69-70 deletion followed by genotyping using SNPsig® SARS-CoV-2 (EscapePLEX) kit (PrimerDesign, UK) that target K417N, E484K, and P681R mutations. The samples with the amplification of spike gene and K417N were termed as probable BA.2 cases. A subset of samples (n=13) were further subjected to whole genome sequencing. The results showed all the 13 samples were of BA.2. Hence, this assay can be used as a cost effective method for the detection of omicron BA.2 variant using real time PCR in resource limiting settings. Moreover, the detection of BA.2 with highly transmissible mutations in Islamabad, Pakistan may potentially increase the number of positive cases. In that scenario, there has to be stringent genomic surveillance to understand the circulating lineages in the country.

## Introduction

The COVID-19 pandemic has witnessed the emergence of several variants of SARS-CoV-2 among which alpha, beta, gamma and delta attained the designation of variants of concern (VOCs). Recently, omicron (lineage B.1.1.529) has been added to the list of VOCs after its detection in South Africa in November 2021. Since its emergence, omicron has spread to all parts of the world leading to new waves of infections in many countries replacing the highly transmissible delta.

Based on genomic diversity, three sub-variants of omicron namely BA.1, BA.2 and BA.3 have been reported. Genome sequencing data from Pakistan shows the circulation of BA.1 (n=42) and BA.1.1 (n=34) as of February 20, 2022. Globally, BA.1 (n=719,364) constitutes the majority of sequences submitted to GISAID followed by BA.1.1 (n=410,015), BA.2 (n=70,570) and BA.3 (n=360) as of February 16, 2022. The BA.2 which differs from BA.1 by approximately 40 mutations [1] has spread to 70 countries since in first detection in South Africa during November 2021. This sub-variant of omicron has become the predominant form in Denmark and is being reportedly increasingly in India, Philippines and Nepal.

Notably, BA.2 lacks the 69-70 deletion of the spike that has given it the name “stealth variant’ due to difficulty in detection using specific real-time PCR kits that target this region. On the contrary, BA.1 that harbors 69-70 deletion can be screened in spike gene target failure samples using specific real-time PCR kits. The Department of Virology, National Institute of Health, Pakistan uses COVID-19 kit (ThermoFisher Scientific) that targets the 69-70 deletion. We used a real-time PCR based algorithm for the detection of BA.2 that is fast, economic and can be used by labs having limited genetic sequencing resources. We confirmed our PCR results through whole-genome sequencing. Moreover, our data shows an increase in BA.2 cases that will provide timely information to the policy makers to take appropriate actions as the country plans to relax COVID-19 restrictions.

## Materials and Methods

### Sampling

Between January 3, 2022, and February 2, 2022, the oropharyngeal swab specimens of suspected subjects were collected as part of routine surveillance at the National Institute of Health’s Department of Virology in Islamabad.

### RNA Extraction and real-time PCR

RNA extraction was performed using the MagMAX Viral/Pathogen Nucleic Acid Isolation kit (ThermoFisher Scientific, USA) and the KingFisher Flex instrument (ThermoFisher Scientific, USA). Clinical RT-PCR testing was performed using the TaqPathTM COVID-19 RT-PCR kit (ThermoFisher Scientific, USA) on an Applied Biosystems 7500 Real-Time PCR system (Thermo Fisher Scientific, USA). The three genes (ORF1ab, N, and S) were targeted. On the basis of spike gene amplification, the samples were divided into two categories: spike gene target failure (SGTF) and non-SGTF. All the samples were further subjected to genotyping using the SNPsig® SARS-CoV-2 (EscapePLEX) kit (PrimerDesign, UK) that target K417N, E484K, and P681R mutations. A subset of non-SGTF samples amplification of K417N were selected for whole genome sequencing.

### cDNA synthesis, and Amplification

The cDNA synthesis and amplification were carried out in accordance with the ARTIC amplicon sequencing protocol (version 2), using SuperScript™ IV VILO™ Master Mix (Invitrogen, USA) and Q5® High-Fidelity 2X Master Mix (New England BioLabs, USA), in combination with the ARTIC nCoV-2019 Panel V3 (Integrated DNA Technologies, Inc, USA) [1].

### Next Generation Sequencing

The Illumina DNA Prep Kit (Illumina, Inc, USA) was used to prepare the paired-end sequencing library (2×150 bp) from the generated amplicons as per manufacturers’ instructions. The prepared libraries were pooled and sequenced on the Illumina iSeq platform with the iSeq 100 i1 Reagent v2 (300-cycle) (Illumina, Inc, USA) at the Department of Virology, National Institute of Health, Islamabad, Pakistan.

### Data Analysis

FastQC (v0.11.9) was used to process the Fastq files for quality assessment [2]. Trimmomatic (v0.39) [3] was used to remove adapter sequences and low-quality base calls (< 30). Alignment of the filtered reads was performed with the available reference genome (Wuhan-Hu-1, GISAID ID: EPI_ISL_402125) using the Burrows-Wheeler Aligner (BWA, v0.7.17) [4]. Variants were identified and consensus sequences for all genomes were generated according to Centers for Disease Control and Prevention (CDC, USA) guidelines [5]. The assembled genomes were classified into PANGO lineages using Pangolin v3.1.19 and the pangoLEARN model dated 20-01-2022 [6].

## Results

During the study period, a total of 558 samples tested positive for SARS-CoV-2 on real-time PCR using the TaqPath™ COVID-19 kit (ThermoFisher Scientific, USA). Among these, 72 (12.9%) samples showed the amplification of all three genes (ORF, N and S), whereas 486 (83.8%) samples had the spike gene target failure (SGTF) i.e. 69-70 deletion. The SARS-CoV-2 genotyping assay (SNPsig EscapePLEX) showed that all samples (n=558) had the K417N mutation whereas E484K, K417T and P681R mutations were not detected (Figure 1). The samples with SGTF (69-70 deletion) and K417N were termed as probable BA.1 while samples with non-SGTF and K417N as probable BA.2. We used this algorithm for the discrimination of omicron sub-lineages BA.1 and BA.2 based on the fact that 69-70 deletion along with K417N mutation is found in omicron BA.1 only whereas K417N alone is characteristic mutation of beta and omicron BA.2. As data from GISAID shows that beta variant is no more prevalent therefore the detection of K417N and absence of E484K, K417T and P681R can be used as a screening method for BA.2. This finding was also confirmed through whole-genome sequencing where a subset of probable BA.2 samples (n=13/13; 100%) were found to be BA.2.

**Figure 1:**
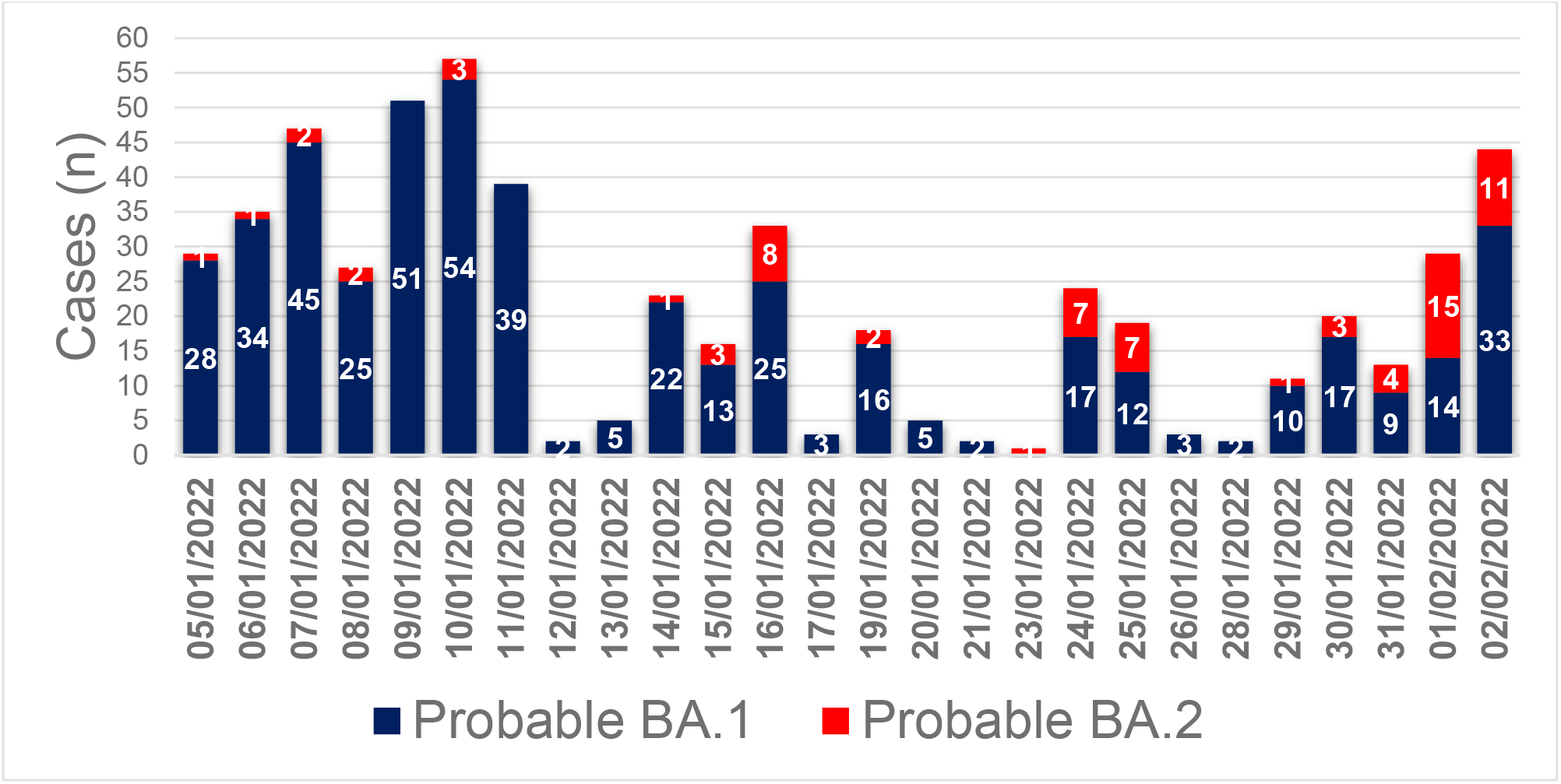
The genotyping data of COVID-19 positive cases during January 5, to February 2, 2022. The SGTF samples with K417N amplification referred as probable BA.1 while non-SGTF with K417N amplification referred as probable BA.2.

Among the thirteen BA.2 cases one patient had a travel history of Turkey while the remaining 12 patients were local resident of Islamabad. Vaccination data revealed, 09 (69%) patients to be fully vaccinated whereas 04 (31%) were unvaccinated. All the isolates exhibited characteristic BA.2 mutations in the ORF1ab (ORF1a:S135R, ORF1a:T842I, ORF1a:G1307S, ORF1a:L3027F, ORF1a:T3090I, ORF1a:L3201F, ORF1a:T3255I, ORF1a:P3395H, ORF1b:P314L, ORF1b:R1315C, ORF1b:I1566V, ORF1b:T2163I, ORF1a:S3675/3677del), ORF3a (T223I), ORF6 (D61L), ORF9b (P10S, E27/29del), membrane (Q19E, A63T), nucleoprotein (P13L, R203K, G204R, S413R, E31/33del) and envelope (T9I) protein. In the spike protein a total of 27 mutations were identified **(Table 1)**. In comparison with the reference BA.2, the study isolates did not exhibit the Y505H mutation.

**Table 1:**
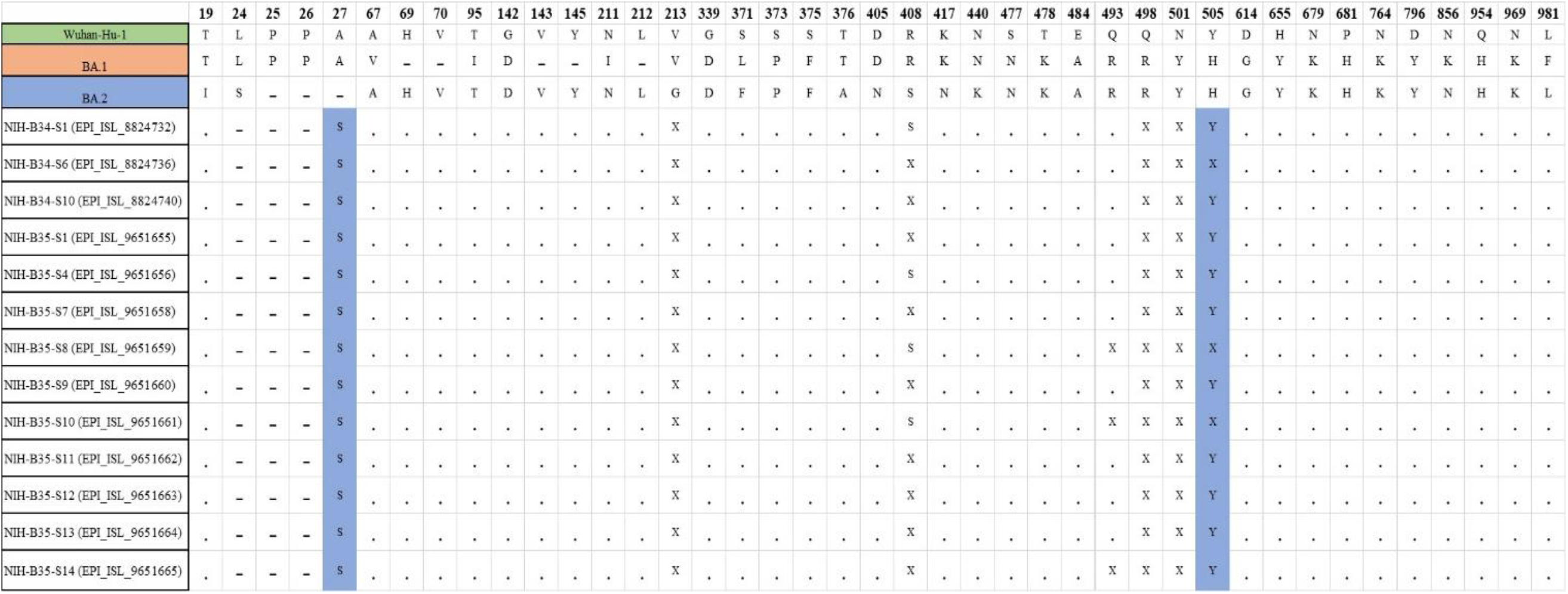
Comparison of mutation profile of spike glycoprotein of SARS-CoV-2 of study isolates with BA.1 and BA.2. The Wuhan-Hu-1 was used as reference. The difference in the mutations of study isolates compared to BA.2 are highlighted in blue color.

## Discussion

The current study identified 72 probable and 13 confirmed cases of omicron sub- lineage BA.2 from Islamabad region of Pakistan with an increasing trend in cases. Although genome sequencing has played a central role in the detection of SARS-CoV-2 variants, monitoring their spread and in control of the disease, however, it remains limited to specialized laboratories that have next generation sequencing equipment, expertise and resources. This has resulted in over representation of SARS-CoV-2 genomic data by such facilities. We used a combination of PCR based genotyping kits as a tool for screening of BA.2 which provides an economical solution for monitoring the spread of this variant in resource limited settings.

After the identification of first case of BA.1 from Pakistan, an increase in number of COVID-19 cases has been observed resulting in fifth wave with cumulative positivity ratio peaking to 12% (as of January 29, 2022). The first case of BA.2 was reported on November 15, 2021, however, in January the number of cases started to rise globally and as of February 20, 2022 the variant has spread to 70 countries. The variant has now become the dominant variant in Denmark (35%), Philippines (78%), Nepal (78%), India (27%) and Singapore (25%) [2]. The variant has also shown the signs of growth in Germany and United Kingdom. Interestingly, the increasing trend in number of subvariant BA.2 cases also suggest that it is increasing relatively more in proportion than the subvariant BA.1. The real world data from these countries suggest that the hospitalization rate, severity of disease, disease symptoms and breakthrough infections are same in case of BA.1 and BA.2 [3-4]. However, a recent study conducted by Yamasoba et al., 2022 has explained that BA.2 is having different virological characteristics than BA.1. It is more pathogenic with reproduction number is 1.4-fold higher than that BA.1. BA.2 has different antigenic profile than BA.1 and it is more fusogenic than BA.1 [5]. The difference in properties of BA.2 and BA.1 makes BA.2 a major risk for global health. The detection of BA.2 in the country has also showed an alarming signal to increase the genomic surveillance and rigorous follow up to understand the circulating lineages in the country. This will also help the policy makers to devise strategies in order to contain the further pandemic waves.

## Data Availability

All data produced are available online at https://www.gisaid.org/.

## References

1. https://fortune.com/2022/01/21/what-is-stealth-omicron-new-covid-variant-substrain-denmark/

2. https://outbreak.info/situationreports?pango=BA.2&loc=IND&selected=Worldwide&overlay=false

3. https://uk.news.yahoo.com/government-monitoring-new-potentially-more-transmissible-omicron-variant-165522269.html

4. https://nyheder.tv2.dk/2022-01-21-ny-omikron-variant-tager-over-i-danmark-det-ved-vi-om-den

5. Yamasoba D, Kimura I, Nasser H, Morioka Y et al. Virological characteristics of SARS-CoV-2 BA.2 variant. 2022. bioRxiv preprint doi: https://doi.org/10.1101/2022.02.14.480335.

